# The emergence, surge and subsequent wave of the SARS-CoV-2 pandemic in New York metropolitan area: The view from a major region-wide urgent care provider

**DOI:** 10.1101/2021.04.06.21255009

**Authors:** Madhura S. Rane, Angela Profeta, Emily Poehlein, Sarah Kulkarni, McKaylee Robertson, Chris Gainus, Ashish Parikh, Kerry LeBenger, Daniel Frogel, Denis Nash

## Abstract

**Background:** Describing SARS-CoV-2 testing and positivity trends among urgent care users is crucial for understanding the trajectory of the pandemic.

**Objective:** To describe demographic and clinical characteristics, positivity rates, and repeat testing patterns among patients tested for SARS-CoV-2 at CityMD, an urgent care provider in the New York City metropolitan area.

**Design:** Retrospective study of all persons testing for SARS-CoV-2 between March 1, 2020 and January 8, 2021 at 115 CityMD locations in the New York metropolitan area.

**Patients:** Individuals receiving a SARS-CoV-2 diagnostic or serologic test.

**Measurements:** Test and individual level SARS-CoV-2 positivity by PCR, rapid antigen, or serologic tests.

**Results:** During the study period, 3.4 million COVID tests were performed on 1.8 million individuals. In New York City, CityMD diagnosed 268,298 individuals, including 17% of all reported cases. Testing levels were higher among 20-29 year olds, non-Hispanic Whites, and females compared with other groups. About 24.8% (n=464,902) were repeat testers. Test positivity was higher in non-Hispanic Black (6.4%), Hispanic (8.0%), and Native American (8.0%) patients compared to non-Hispanic White (5.4%) patients. Overall seropositivity was estimated to be 21.7% (95% Confidence Interval [CI]: 21.6-21.8) and was highest among 10-14 year olds (27.3%). Seropositivity was also high among non-Hispanic Black (24.5%) and Hispanic (30.6%) testers, and residents of the Bronx (31.3%) and Queens (30.5%). Using PCR as the gold standard, SARS-CoV-2 rapid tests had a false positive rate of 5.4% (95%CI 5.3-5.5).

**Conclusion:** Urgent care centers can provide broad access to critical evaluation, diagnostic testing and treatment of a substantial number of ambulatory patients during pandemics, especially in population-dense, urban epicenters.

## Introduction

New York City (NYC) was the epicenter of the COVID-19 pandemic in March 2020. In the beginning of the pandemic, the COVID-19 testing criteria in New York (NY) State were highly restrictive due to limited availability of tests [1], with only severe and hospitalized cases being tested. On March 7, NY’s governor declared a state of emergency, which allowed for expedited purchasing and an expanded testing protocol that covered patients without an identified exposure but experiencing severe symptoms. Commercial laboratories also began testing for SARS-CoV-2. By June 2, anyone in NY could be tested regardless of symptoms or exposure. Repeat testing was recommended for those who worked in residential congregate settings or who had ongoing concerns around possible SARS-CoV-2 exposure [2].

Widespread availability of diagnostic tests for SARS-CoV-2, including polymerase chain reaction (PCR) and point-of-care rapid antigen tests, is instrumental in limiting transmission when test turnaround time is fast and positive tests are followed by self-isolation [3]. Urgent care providers can play a crucial role in meeting the large demand for COVID-19 clinical evaluation and testing by providing immediate access for symptomatic patients and those with high-risk exposures, thus reducing unnecessary use of hospital emergency departments. Additionally, urgent care centers provide prompt access to patients who have mild symptoms, who are asymptomatic, or who require repeat testing when such access at doctor’s offices is limited.

Describing SARS-CoV2 testing patterns among urgent care patients and demographic and clinical characteristics of those who tested positive can provide key insights into the trajectory of the pandemic. Data on repeat testing in the general population is also lacking. Finally, operating characteristics for rapid antigen tests, increasingly used in routine practice settings, have not been well-characterized at scale. We describe SARS-CoV-2 testing and testing outcomes during the SARS-CoV-2 pandemic in the New York metropolitan area using electronic medical records (EMRs) from a major urgent care provider.

## Methods

### Study setting and participants

This study includes all patients who received a COVID-19 diagnostic or serologic test at CityMD urgent care sites in the New York metropolitan area (NYC, Long Island and Westchester areas). Pending or missing test results were excluded. CityMD is the largest walk-in urgent medical care provider in the region and was a frontline provider for COVID-19 diagnostics and treatment at the earliest phase of the pandemic. It has since been serving as one the area’s largest COVID-19 clinical evaluation and SARS-CoV-2 testing providers.

### Data collection

Two types of COVID-19 diagnostic tests were offered at CityMD: PCR tests and rapid antigen tests. Serologic testing was also offered. We examined de-identified EMR data and all SARS-CoV-2 diagnostic and serological test results between March 1, 2020 - January 8, 2021 from CityMD’s 115 locations in the five boroughs of NYC (n=76), and the surrounding suburban areas of Long Island (n=32) and Westchester (n=7). Testing, using assays authorized for emergency use by the Food and Drug Administration (FDA), included: 1) PCR tests of respiratory tract specimens for SARS-CoV-2 RNA collected via nasopharyngeal and nasal swabs; 2) serologic tests of serum specimens, and 3) rapid antigen tests of respiratory tract specimens collected via anterior nasal swabs. PCR and serologic tests were conducted by commercial laboratories, and rapid antigen tests were conducted on-site.

All patients were evaluated by a licensed clinician. We examined routinely collected data on body temperature and oxygen saturation at the time of COVID-19 testing. Additionally, we examined the presence of COVID-19 symptoms among patients who received rapid tests.

### Demographic characteristics

Individual-level demographic factors such as age, gender, race, ethnicity, and region of residence at the time of testing were examined. Age at time of visit was categorized into 5 year intervals up to 20 years, and in 10 year intervals for those 20 years and older, going up to >100 years.Self-reported race and ethnicity data was mapped to the US Office of Management and Budget (OMB) defined categories for race/ethnicity [4] (See Supplement for detailed breakdowns).

### Definitions

Test-level positivity for PCR and rapid antigen tests was defined as the percent of total tests performed on a given day that were positive. Individual-level percent positivity is defined as the percent of individuals tested on a given day with a positive result. For individuals with PCR and rapid antigen tests on the same day, we used the PCR test result in estimating the daily test positivity. For individuals with more than one positive test on different days, only the first positive result was included for estimating daily individual-level positivity rate trend. We plotted the cumulative number of individuals with *any* positive SARS-CoV-2 diagnostic or serologic test (PCR, antigen, antibody) during the study period to assess the total number of individuals with evidence of current or prior SARS-CoV-2 infection at the time of clinic visit.

Repeat testing was defined as having two or more diagnostic tests (PCR or antigen) on separate days.

For individuals who had a confirmatory PCR test following a rapid antigen test, we estimated the false negative and false positive rate of the rapid test, with PCR test result from the same specimen as the gold standard.

### Statistical analysis

Descriptive statistics were used to summarize COVID-19 testing and test positivity by demographic characteristics at the test and individual levels. We compared COVID-19 positivity trends in three time periods: from March to June (emergence and wave 1), July to September (low activity), and October to January 2021 (wave 2). We plotted the daily test volume, proportion of positive tests, and proportion of individuals testing positive for PCR, rapid antigen, and serologic tests to assess temporal trends. All analyses were conducted in R v4.0.1.

### Ethical Review

This study was approved by the Institutional Review Board of the City University of New York Graduate School of Public Health and Health Policy.

## Results

Between March 1, 2020 and January 8, 2021, CityMD performed 3.4 million diagnostic and serologic tests on 1.8 million individuals living in NYC and surrounding areas of Long Island and Westchester. We excluded tests with pending (n=84,648), missing (n=8,447), or unmapped (n=494) results (2.7% of tests). Testers were commonly 20-29 years old, non-Hispanic White, and female (Table 1). Overall, 43% of the total tests (n=1,489,960) performed were PCR tests, 35% (n=1,211,957) were rapid antigen tests, and 22% (n=752,643) were serological tests. Prior to October, 59% (n=1,005,145) of all tests performed were PCR tests and 41% (n=697,911) were serological tests. Since their introduction in October, the majority of the tests performed have been rapid antigen tests (n=1,211,957 [69.2%]). The usage of serologic tests decreased as the epidemic progressed. During the study period, CityMD diagnosed 268,298 individuals with SARS-CoV-2 infection, of which 183,970 patients were seen in NYC (sFig 1).

**Table 1:**
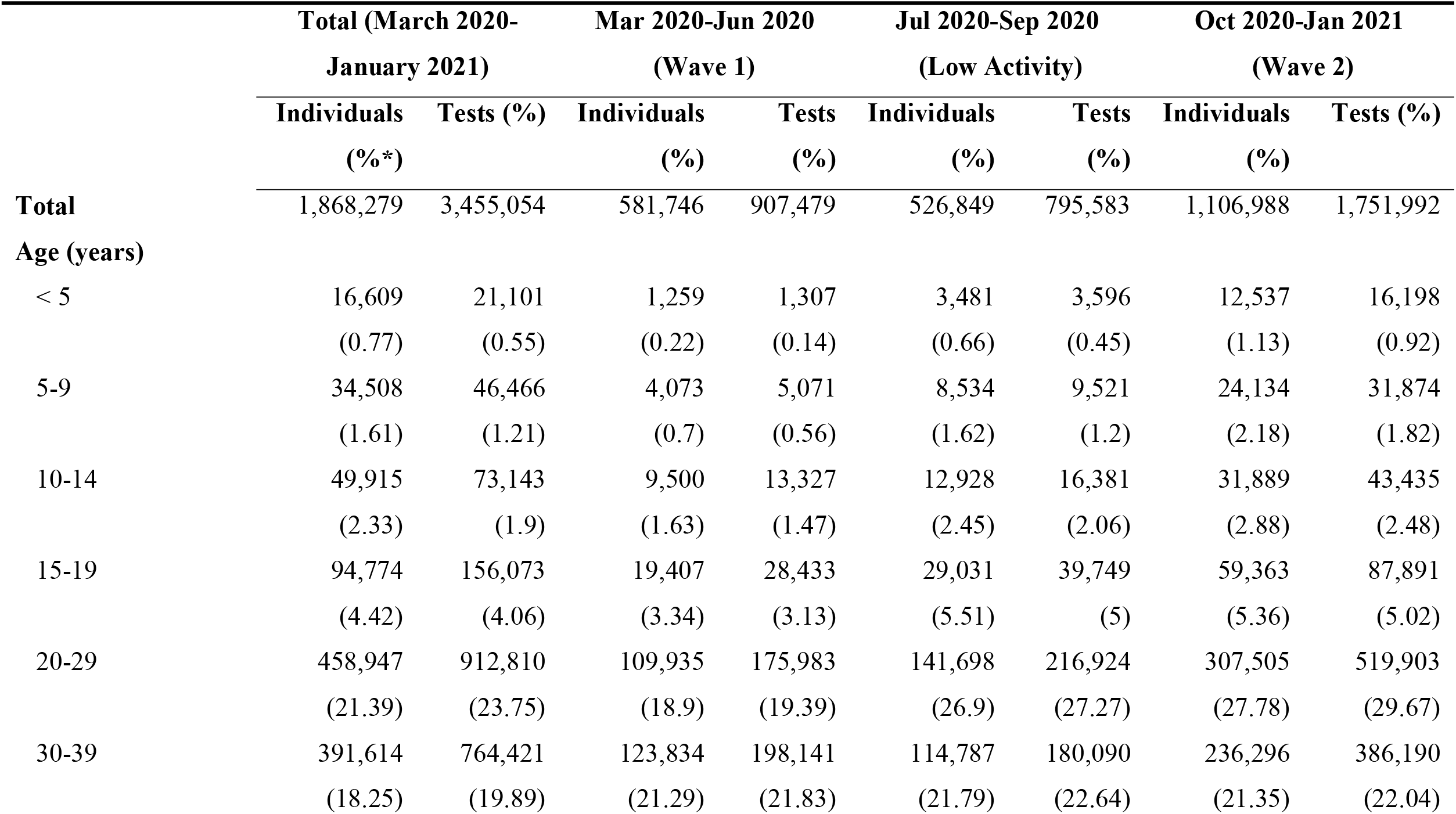
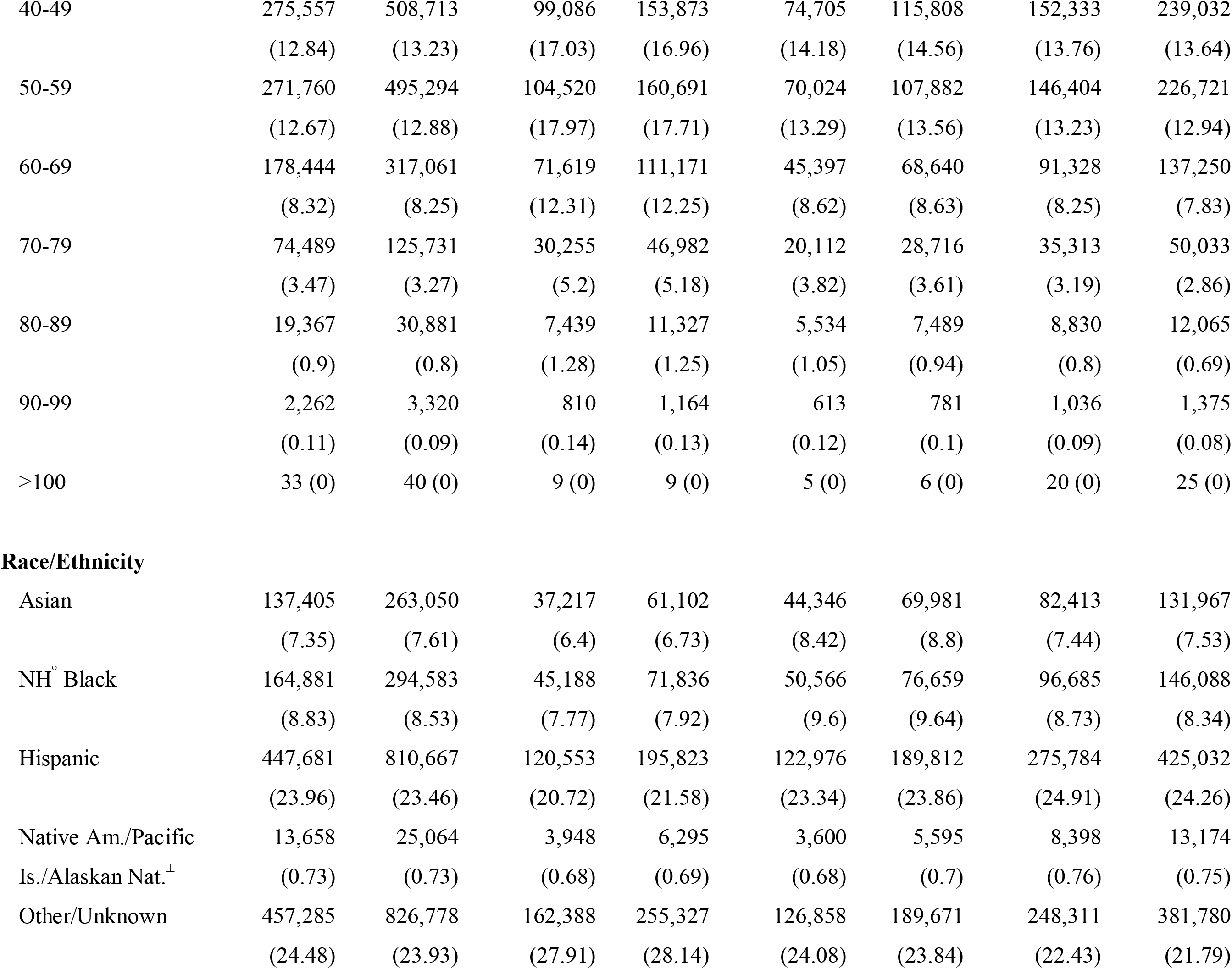
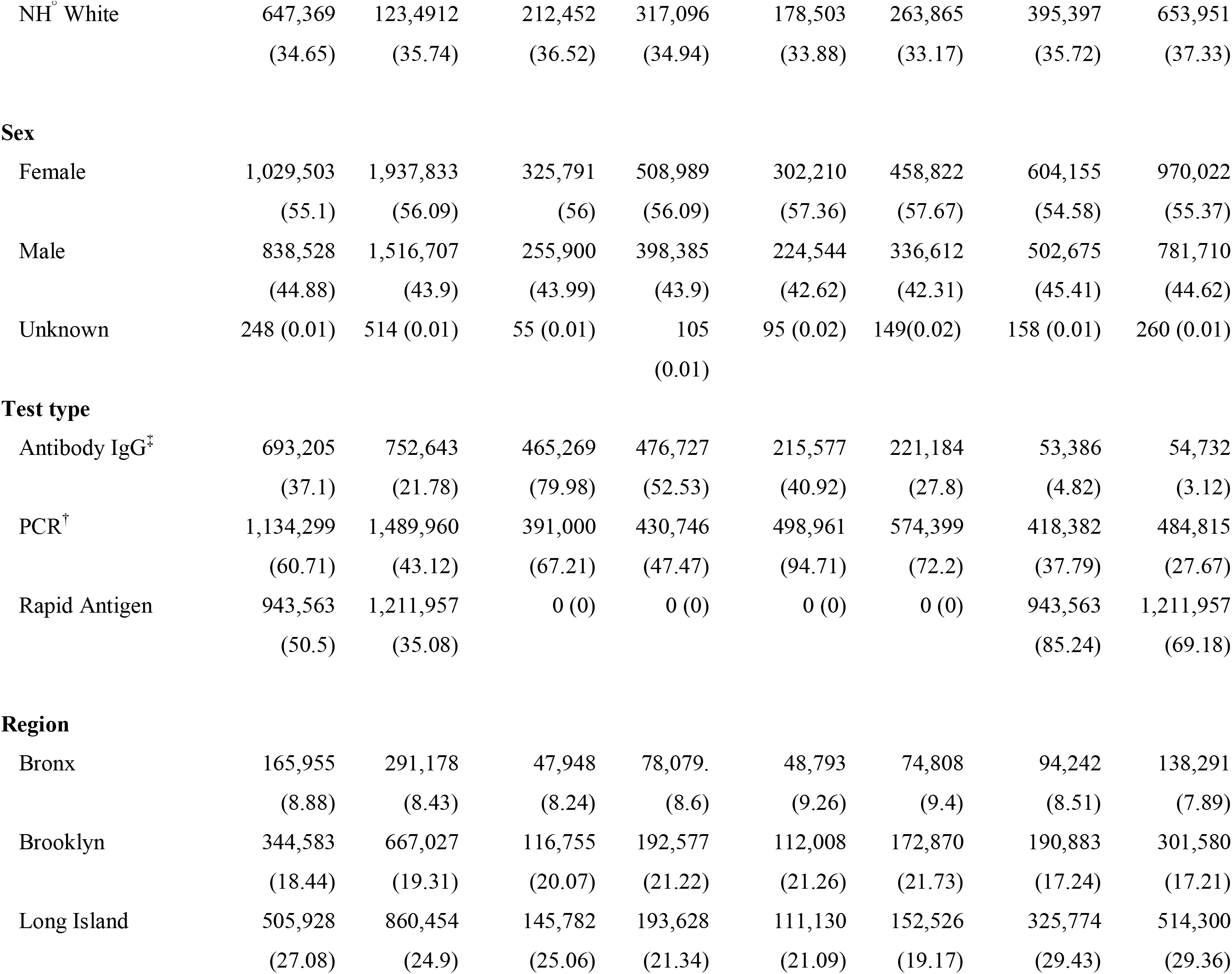
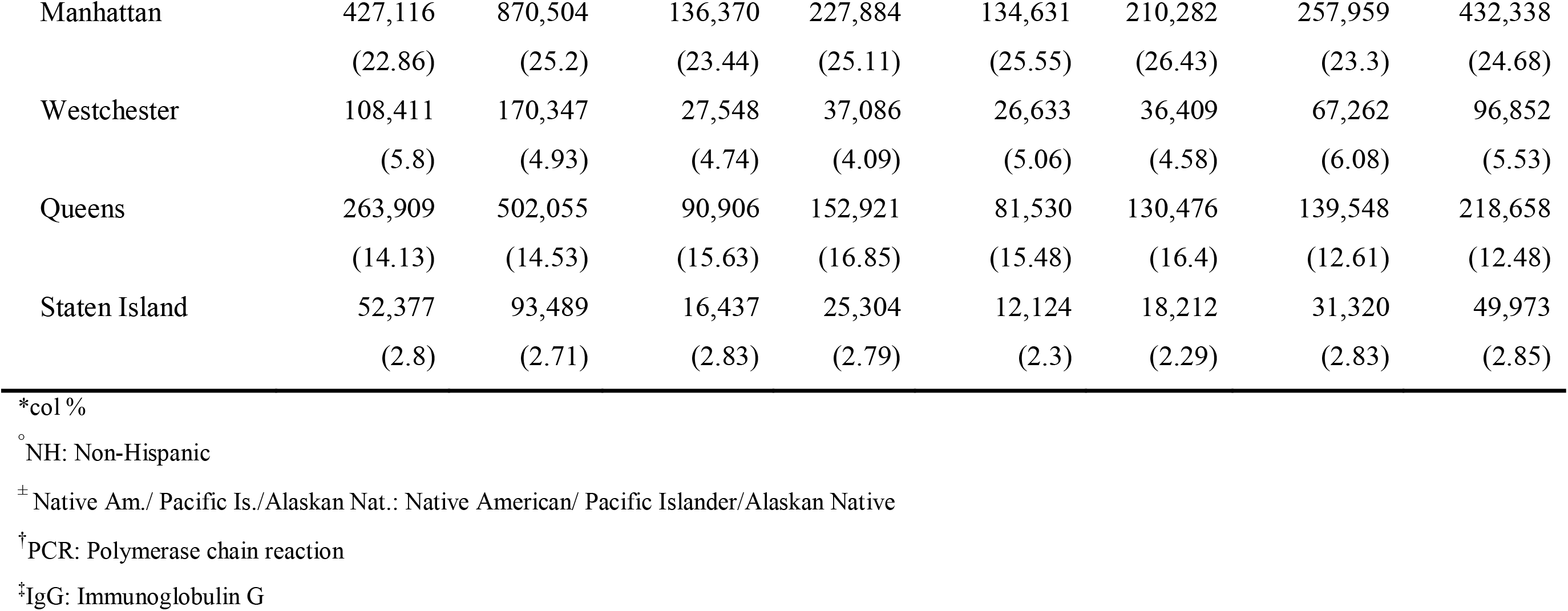
Demographic characteristics of patients tested for SARS-CoV-2 by PCR, antigen, and serology tests at CityMD, March 1, 2020-January 8, 2021.

|  | Total (March 2020-January 2021) |  | Mar 2020-Jun 2020 (Wave 1) |  | Jul 2020-Sep 2020 (Low Activity) |  | Oct 2020-Jan 2021 (Wave 2) |  |
| --- | --- | --- | --- | --- | --- | --- | --- | --- |
|  | Individuals | Tests (%) | Individuals | Tests | Individuals | Tests | Individuals | Tests (%) |
|  | (%*) |  | (%) | (%) | (%) | (%) | (%) |  |
| **Total** | 1,868,279 | 3,455,054 | 581,746 | 907,479 | 526,849 | 795,583 | 1,106,988 | 1,751,992 |
| **Age (years)** |  |  |  |  |  |  |  |  |
| < 5 | 16,609 | 21,101 | 1,259 | 1,307 | 3,481 | 3,596 | 12,537 | 16,198 |
|  | (0.77) | (0.55) | (0.22) | (0.14) | (0.66) | (0.45) | (1.13) | (0.92) |
| 5-9 | 34,508 | 46,466 | 4,073 | 5,071 | 8,534 | 9,521 | 24,134 | 31,874 |
|  | (1.61) | (1.21) | (0.7) | (0.56) | (1.62) | (1.2) | (2.18) | (1.82) |
| 10-14 | 49,915 | 73,143 | 9,500 | 13,327 | 12,928 | 16,381 | 31,889 | 43,435 |
|  | (2.33) | (1.9) | (1.63) | (1.47) | (2.45) | (2.06) | (2.88) | (2.48) |
| 15-19 | 94,774 | 156,073 | 19,407 | 28,433 | 29,031 | 39,749 | 59,363 | 87,891 |
|  | (4.42) | (4.06) | (3.34) | (3.13) | (5.51) | (5) | (5.36) | (5.02) |
| 20-29 | 458,947 | 912,810 | 109,935 | 175,983 | 141,698 | 216,924 | 307,505 | 519,903 |
|  | (21.39) | (23.75) | (18.9) | (19.39) | (26.9) | (27.27) | (27.78) | (29.67) |
| 30-39 | 391,614 | 764,421 | 123,834 | 198,141 | 114,787 | 180,090 | 236,296 | 386,190 |
|  | (18.25) | (19.89) | (21.29) | (21.83) | (21.79) | (22.64) | (21.35) | (22.04) |
| 40-49 | 275,557<br>(12.84) | 508,713<br>(13.23) | 99,086<br>(17.03) | 153,873<br>(16.96) | 74,705<br>(14.18) | 115,808<br>(14.56) | 152,333<br>(13.76) | 239,032<br>(13.64) |
| 50-59 | 271,760<br>(12.67) | 495,294<br>(12.88) | 104,520<br>(17.97) | 160,691<br>(17.71) | 70,024<br>(13.29) | 107,882<br>(13.56) | 146,404<br>(13.23) | 226,721<br>(12.94) |
| 60-69 | 178,444<br>(8.32) | 317,061<br>(8.25) | 71,619<br>(12.31) | 111,171<br>(12.25) | 45,397<br>(8.62) | 68,640<br>(8.63) | 91,328<br>(8.25) | 137,250<br>(7.83) |
| 70-79 | 74,489<br>(3.47) | 125,731<br>(3.27) | 30,255<br>(5.2) | 46,982<br>(5.18) | 20,112<br>(3.82) | 28,716<br>(3.61) | 35,313<br>(3.19) | 50,033<br>(2.86) |
| 80-89 | 19,367<br>(0.9) | 30,881<br>(0.8) | 7,439<br>(1.28) | 11,327<br>(1.25) | 5,534<br>(1.05) | 7,489<br>(0.94) | 8,830<br>(0.8) | 12,065<br>(0.69) |
| 90-99 | 2,262<br>(0.11) | 3,320<br>(0.09) | 810<br>(0.14) | 1,164<br>(0.13) | 613<br>(0.12) | 781<br>(0.1) | 1,036<br>(0.09) | 1,375<br>(0.08) |
| >100 | 33 (0) | 40 (0) | 9 (0) | 9 (0) | 5 (0) | 6 (0) | 20 (0) | 25 (0) |
| **Race/Ethnicity** |  |  |  |  |  |  |  |  |
| Asian | 137,405<br>(7.35) | 263,050<br>(7.61) | 37,217<br>(6.4) | 61,102<br>(6.73) | 44,346<br>(8.42) | 69,981<br>(8.8) | 82,413<br>(7.44) | 131,967<br>(7.53) |
| NH<sup>o</sup> Black | 164,881<br>(8.83) | 294,583<br>(8.53) | 45,188<br>(7.77) | 71,836<br>(7.92) | 50,566<br>(9.6) | 76,659<br>(9.64) | 96,685<br>(8.73) | 146,088<br>(8.34) |
| Hispanic | 447,681<br>(23.96) | 810,667<br>(23.46) | 120,553<br>(20.72) | 195,823<br>(21.58) | 122,976<br>(23.34) | 189,812<br>(23.86) | 275,784<br>(24.91) | 425,032<br>(24.26) |
| Native Am./Pacific<br>Is./Alaskan Nat.<sup>±</sup> | 13,658<br>(0.73) | 25,064<br>(0.73) | 3,948<br>(0.68) | 6,295<br>(0.69) | 3,600<br>(0.68) | 5,595<br>(0.7) | 8,398<br>(0.76) | 13,174<br>(0.75) |
| Other/Unknown | 457,285<br>(24.48) | 826,778<br>(23.93) | 162,388<br>(27.91) | 255,327<br>(28.14) | 126,858<br>(24.08) | 189,671<br>(23.84) | 248,311<br>(22.43) | 381,780<br>(21.79) |
| NH° White | 647,369<br>(34.65) | 123,4912<br>(35.74) | 212,452<br>(36.52) | 317,096<br>(34.94) | 178,503<br>(33.88) | 263,865<br>(33.17) | 395,397<br>(35.72) | 653,951<br>(37.33) |
| **Sex** |  |  |  |  |  |  |  |  |
| Female | 1,029,503<br>(55.1) | 1,937,833<br>(56.09) | 325,791<br>(56) | 508,989<br>(56.09) | 302,210<br>(57.36) | 458,822<br>(57.67) | 604,155<br>(54.58) | 970,022<br>(55.37) |
| Male | 838,528<br>(44.88) | 1,516,707<br>(43.9) | 255,900<br>(43.99) | 398,385<br>(43.9) | 224,544<br>(42.62) | 336,612<br>(42.31) | 502,675<br>(45.41) | 781,710<br>(44.62) |
| Unknown | 248 (0.01) | 514 (0.01) | 55 (0.01) | 105<br>(0.01) | 95 (0.02) | 149(0.02) | 158 (0.01) | 260 (0.01) |
| **Test type** |  |  |  |  |  |  |  |  |
| Antibody IgG<sup>‡</sup> | 693,205<br>(37.1) | 752,643<br>(21.78) | 465,269<br>(79.98) | 476,727<br>(52.53) | 215,577<br>(40.92) | 221,184<br>(27.8) | 53,386<br>(4.82) | 54,732<br>(3.12) |
| PCR<sup>†</sup> | 1,134,299<br>(60.71) | 1,489,960<br>(43.12) | 391,000<br>(67.21) | 430,746<br>(47.47) | 498,961<br>(94.71) | 574,399<br>(72.2) | 418,382<br>(37.79) | 484,815<br>(27.67) |
| Rapid Antigen | 943,563<br>(50.5) | 1,211,957<br>(35.08) | 0 (0) | 0 (0) | 0 (0) | 0 (0) | 943,563<br>(85.24) | 1,211,957<br>(69.18) |
| **Region** |  |  |  |  |  |  |  |  |
| Bronx | 165,955<br>(8.88) | 291,178<br>(8.43) | 47,948<br>(8.24) | 78,079.<br>(8.6) | 48,793<br>(9.26) | 74,808<br>(9.4) | 94,242<br>(8.51) | 138,291<br>(7.89) |
| Brooklyn | 344,583<br>(18.44) | 667,027<br>(19.31) | 116,755<br>(20.07) | 192,577<br>(21.22) | 112,008<br>(21.26) | 172,870<br>(21.73) | 190,883<br>(17.24) | 301,580<br>(17.21) |
| Long Island | 505,928<br>(27.08) | 860,454<br>(24.9) | 145,782<br>(25.06) | 193,628<br>(21.34) | 111,130<br>(21.09) | 152,526<br>(19.17) | 325,774<br>(29.43) | 514,300<br>(29.36) |
| Manhattan | 427,116 | 870,504 | 136,370 | 227,884 | 134,631 | 210,282 | 257,959 | 432,338 |
|  | (22.86) | (25.2) | (23.44) | (25.11) | (25.55) | (26.43) | (23.3) | (24.68) |
| Westchester | 108,411 | 170,347 | 27,548 | 37,086 | 26,633 | 36,409 | 67,262 | 96,852 |
|  | (5.8) | (4.93) | (4.74) | (4.09) | (5.06) | (4.58) | (6.08) | (5.53) |
| Queens | 263,909 | 502,055 | 90,906 | 152,921 | 81,530 | 130,476 | 139,548 | 218,658 |
|  | (14.13) | (14.53) | (15.63) | (16.85) | (15.48) | (16.4) | (12.61) | (12.48) |
| Staten Island | 52,377 | 93,489 | 16,437 | 25,304 | 12,124 | 18,212 | 31,320 | 49,973 |
|  | (2.8) | (2.71) | (2.83) | (2.79) | (2.3) | (2.29) | (2.83) | (2.85) |

\*col %
° NH: Non-Hispanic
± Native Am./ Pacific Is./Alaskan Nat.: Native American/ Pacific Islander/Alaskan Native
† PCR: Polymerase chain reaction
‡ IgG: Immunoglobulin G

**Figure 1:**
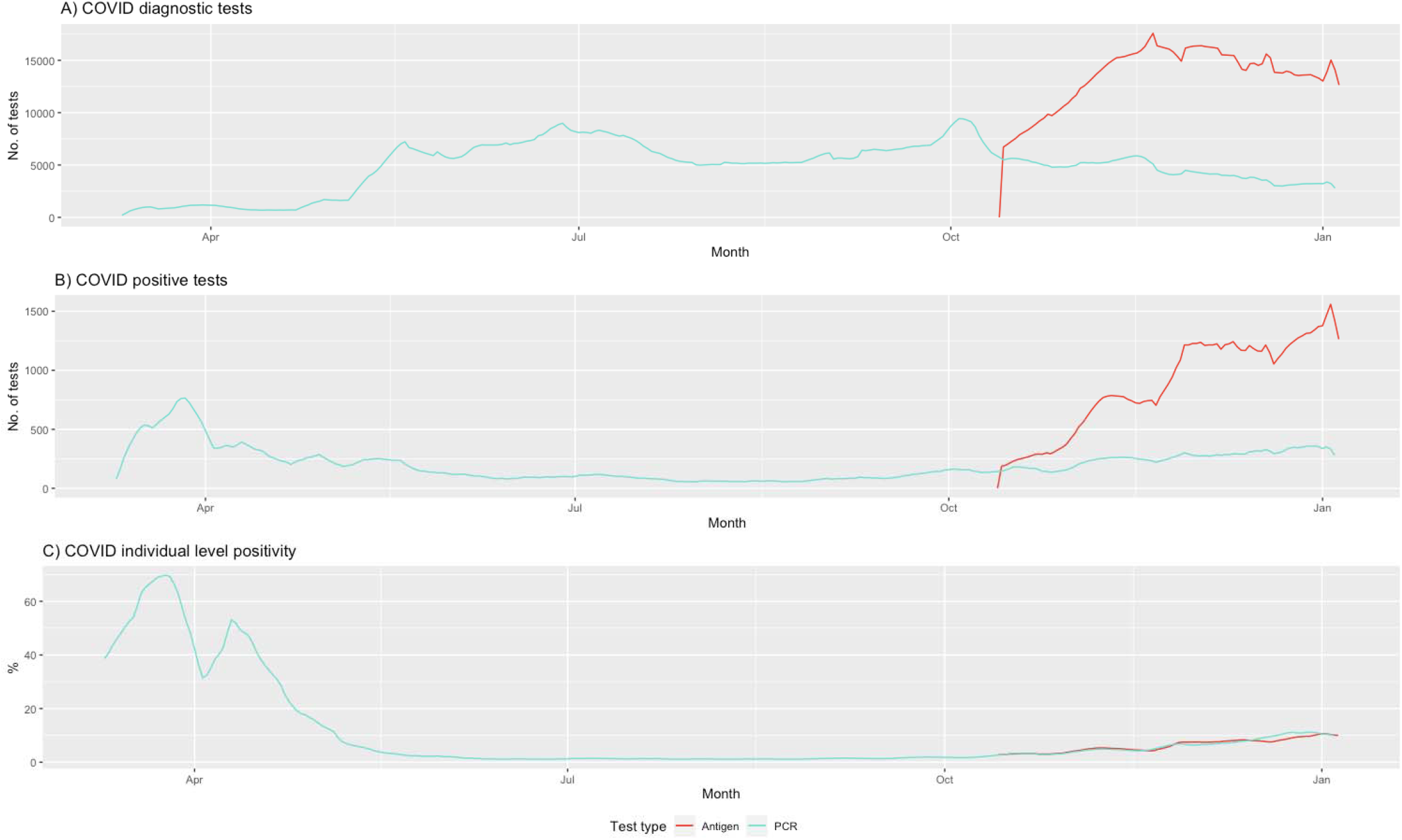
COVID molecular and rapid antigen testing trends at CityMD in the New York metropolitan area. A seven-day rolling average of tests is plotted to smooth temporal trends. PCR tests are denoted in turquoise and rapid antigen tests are in red. Panels A and B show daily tests performed and number of tests that were positive. Panel C shows the proportion of individuals who received their first positive test by PCR and antigen tests over time (individual-level daily positivity rate). Rapid antigen testing was only offered starting October. Note that initial positivity rates were high because tests were in short supply and testing criteria were stringent. PCR: Polymerase chain reaction.

### Testing patterns

Of the 1,868,279 individuals who were tested, 908,889 (48.6%) received ≥2 tests of any type (sFig2), among whom 643,853 (70.8%) received 2 tests *on the same day*. Of these, 442,822 (68.7%) received both a serologic and a PCR test, 212,948 (33.1%) received both a PCR and a rapid test, 29,322 (4.5%) individuals received both a serologic and a rapid test (Tables 2 and 3).

**Table 2:**
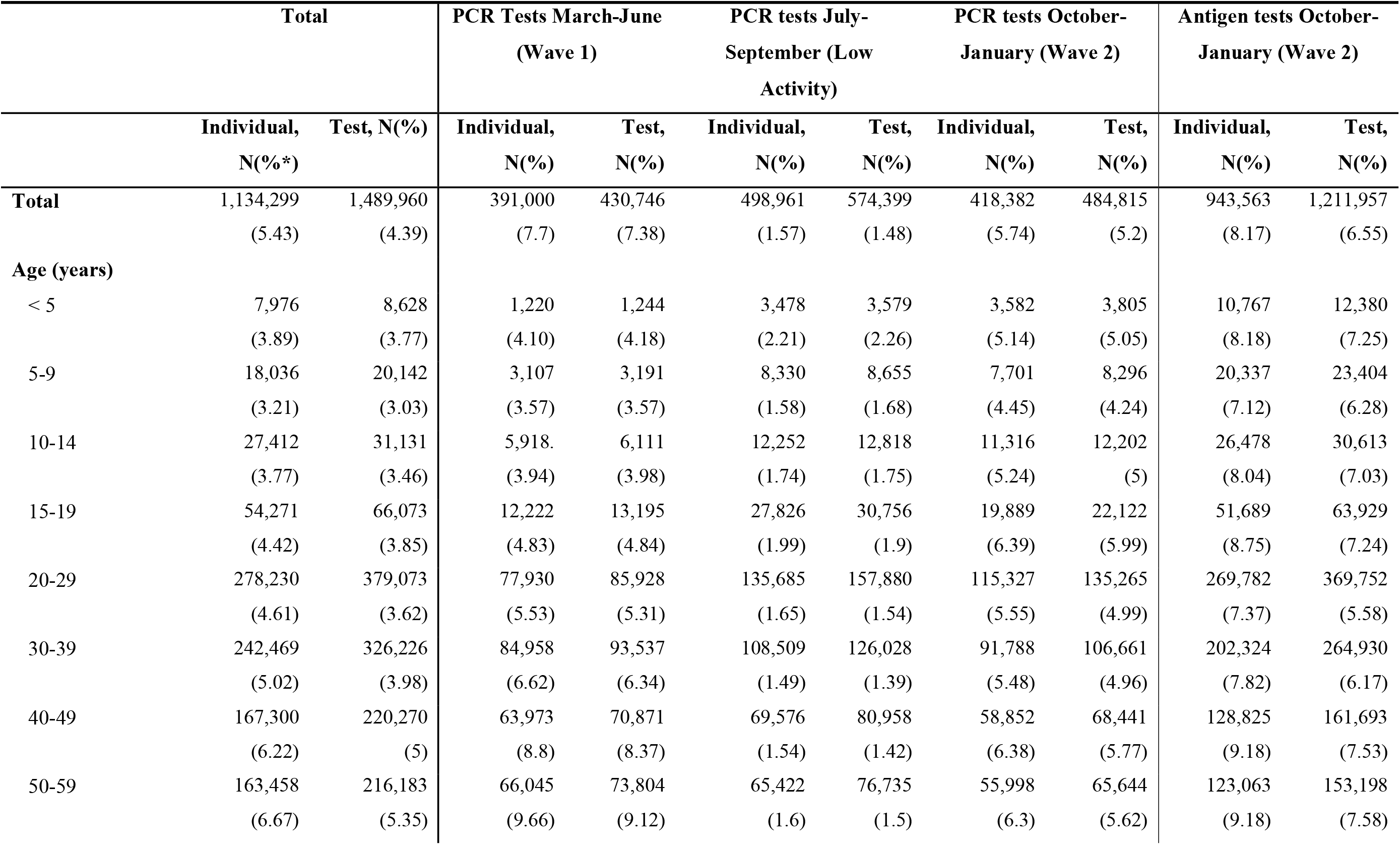
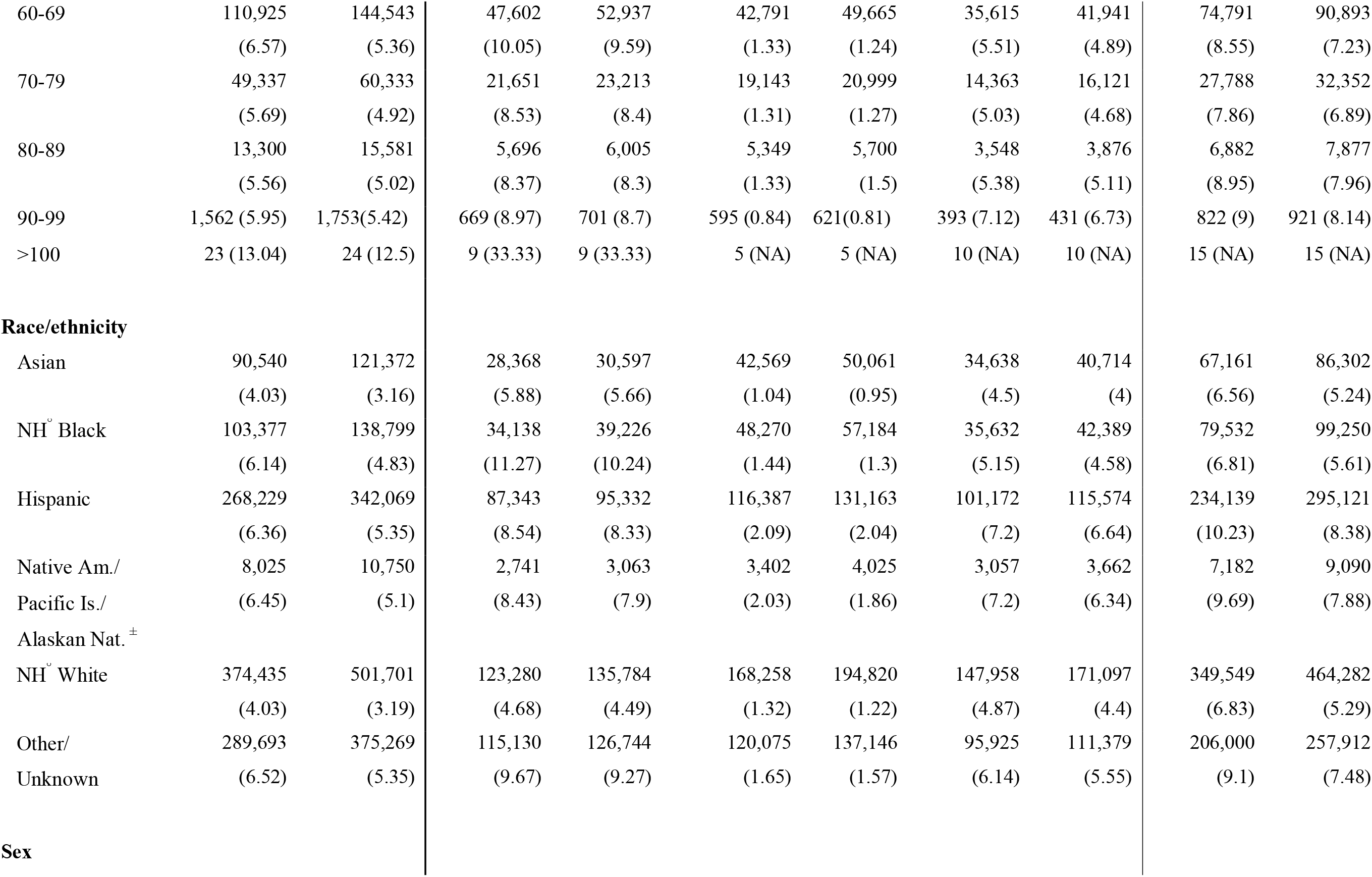
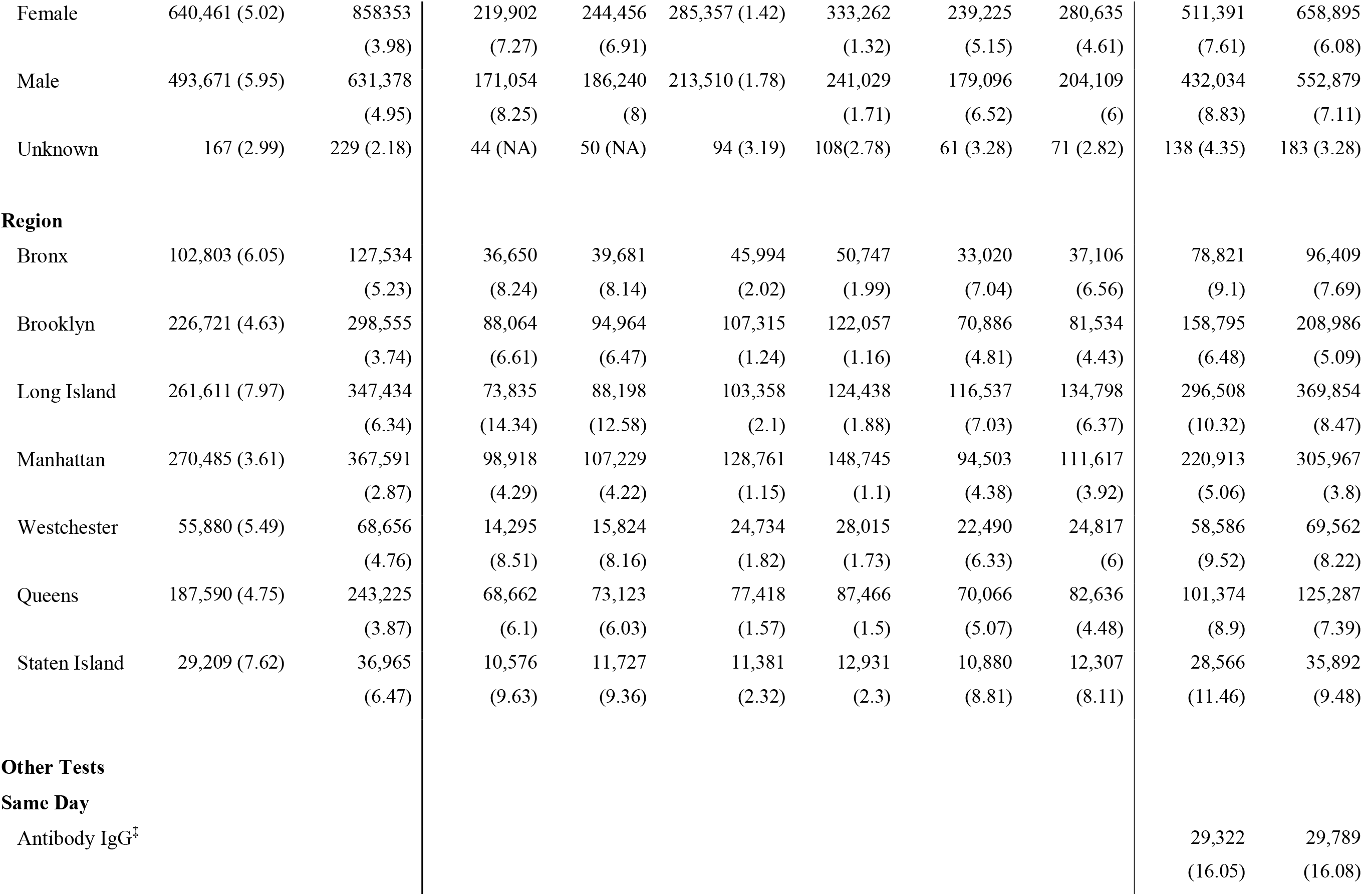
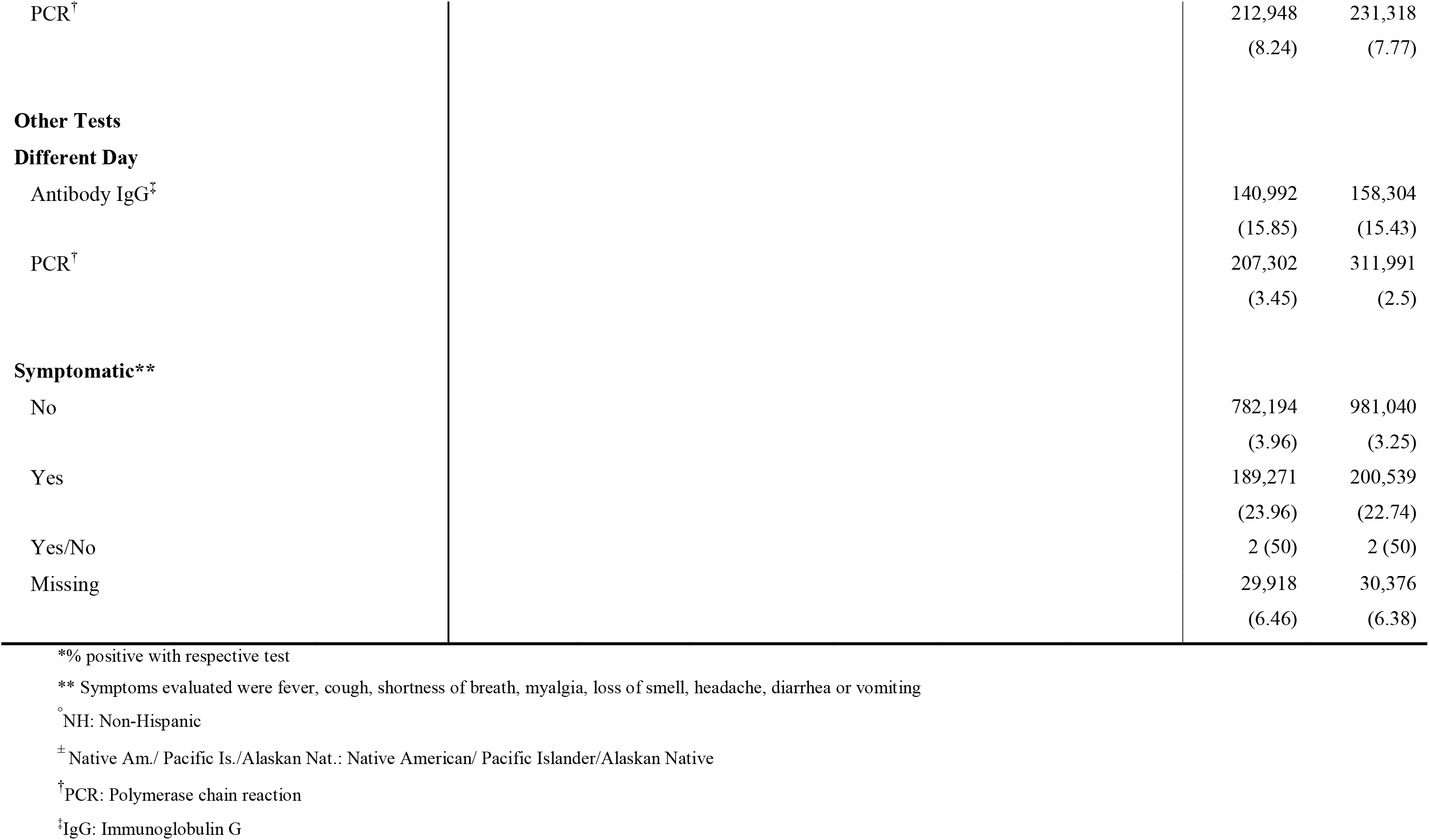
Demographic characteristics of patients tested and diagnosed with SARS-CoV-2 by PCR and antigen test at CityMD, March 1, 2020-January 8, 2021.

**Table 3.** Demographic characteristics of patients who received a SARS-CoV-2 serologic test an tested positive at a large urgent care provider, March 01, 2020-January 8, 2021.

|  | Individual, N (%*) | Total, N (%) |
| --- | --- | --- |
| **Total** | 693,205 (21.66) | 752,643 (21.15) |
| **Age (years)** |  |  |
| < 5 | 91 (16.48) | 93 (16.13) |
| 5-9 | 2,813 (26.91) | 2,920 (26.82) |
| 10-14 | 10,984 (27.26) | 11,396 (26.93) |
| 15-19 | 24,636 (23.51) | 26,058 (22.83) |
| 20-29 | 148,801 (18.18) | 163,802 (17.45) |
| 30-39 | 157,453 (19.2) | 173,119 (18.56) |
| 40-49 | 116,947 (23.62) | 126,667 (23.09) |
| 50-59 | 116,674 (25.01) | 125,872 (24.65) |
| 60-69 | 76,128 (24.25) | 81,605 (24.2) |
| 70-79 | 31,036 (20.82) | 33,041 (21.08) |
| 80-89 | 7,015 (20.29) | 7,423 (20.4) |
| 90-99 | 626 (21.25) | 646 (21.52) |
| >100 | 1 (100) | 1 (100) |
| **Race** |  |  |
| Asian | 50,804 (15.6) | 55,314 (15.41) |
| NH<sup>o</sup> Black | 521,42 (25.45) | 56,495 (24.61) |
| Hispanic | 160,026 (30.58) | 173,358 (29.59) |
| Native Am./ Pacific Is./Alaskan Nat. <sup>±</sup> | 4,792 (24.37) | 5,221 (23.65) |
| Other/Unknown | 178,847 (25.92) | 193,501 (25.42) |
| NH<sup>o</sup> White | 246,594 (13.17) | 268,754 (13.03) |
| **Sex** |  |  |
| Female | 388,897 (21.13) | 420,294 (20.72) |
| Male | 304,218 (22.33) | 332,247 (21.69) |
| Unknown | 90 (12.22) | 102 (10.78) |
| **Other tests same day** |  |  |
| COVID PCR<sup>†</sup> (Active) | 442,822 (2.04) | 475,774 (1.94) |
| Rapid antigen Test | 29,322 (1.63) | 29,783 (1.6) |
| **Other tests different day** |  |  |
| COVID PCR<sup>†</sup> (Active) | 163,930 (6.64) | 250,956 (4.67) |
| Rapid antigen Test | 142,404 (6.17) | 204,282 (4.46) |
| **Region** |  |  |
| NYC (Overall) | 528,230 (22.7) | 577,469 (22.1) |
| Bronx | 62,333 (31.26) | 67,220 (30.37) |
| Brooklyn | 145,917 (19.77) | 159,428 (19.09) |
| Long Island | 134,517 (18.42) | 143,139 (18.23) |
| Manhattan | 177,846 (17.03) | 196,717 (16.67) |
| Metro North | 30,458 (17.78) | 32,035 (17.6) |
| Queens | 123,076 (30.53) | 133,490 (29.84) |
| Staten Island | 19,058 (19.62) | 20,614 (19.28) |

\*% tested positive with serologic test
° NH: Non-Hispanic
± Native Am./ Pacific Is./Alaskan Nat.: Native American/ Pacific Islander/Alaskan Native
† PCR: Polymerase chain reaction

Of the 1.8 million patients tested, 523,502 (28%) received ≥2 diagnostic and/or serologic tests *on separate days*. Of these, the majority had 2 visits (n=348,002, [66.4%]), 107,968 testers (20.6%) had 3 visits, while 64,980 testers (12.4%) had between 4 and 10 visits. A small number of testers (n=3,106 [0.6%] had 10 or more tests. A total of 25,833 (4.9%) repeat testers received their first serologic test after a diagnostic test (median time interval: 34 days; IQR: 15-60 days), while 77,856 (14.9%) received their first diagnostic test after a serologic test (median time interval between first antibody test and first diagnostic test: 121 days; IQR: 56-180 days).

A total of 464,902 (88.8% of those with ≥2 tests) patients had two or more *diagnostic tests* on separate days (repeat testers; median time interval between first and last diagnostic test: 63 days, IQR: (24-134). Most repeat diagnostic testing occurred between October 2020 and January 2021. Of these, 7,303 testers (1.5%) had multiple positive tests (median interval between first and last positive diagnostic tests: 11 days; IQR: 7-15 days). Only 52 of the 7,303 (0.7%) cases had an interval of >90 between two positive tests, i.e. potential re-infections.

### Temporal trends in COVID testing and positivity

Testing increased rapidly after the last week of April and stayed high. The introduction of rapid antigen tests in October 2020 was accompanied by a further steep increase in testing (Fig 1A). The individual-level positivity rate was high early in the pandemic mainly because SARS-CoV-2 testing in New York was restricted to severe or hospitalized cases (Fig 1C).

Trends in the number of positive tests and percent positivity started declining in late March coinciding with the strict lockdown and physical distancing mandates implemented in March. The daily individual-level positivity rate remained low (∼1%) until September. Starting in October, when physical distancing rules were relaxed, and indoor dining, bars, and schools reopened, cases once again increased rapidly giving way to a second wave of the pandemic (Figs 1B and 1C). Daily testing and positivity rates continued to increase through NY’s 2020 federal election early voting period in November and during holidays such as Thanksgiving and Christmas (Fig 2). As of January 8, 2021, diagnostic test positivity rates had climbed to ∼10% (Fig 1B).

**Figure 2:**
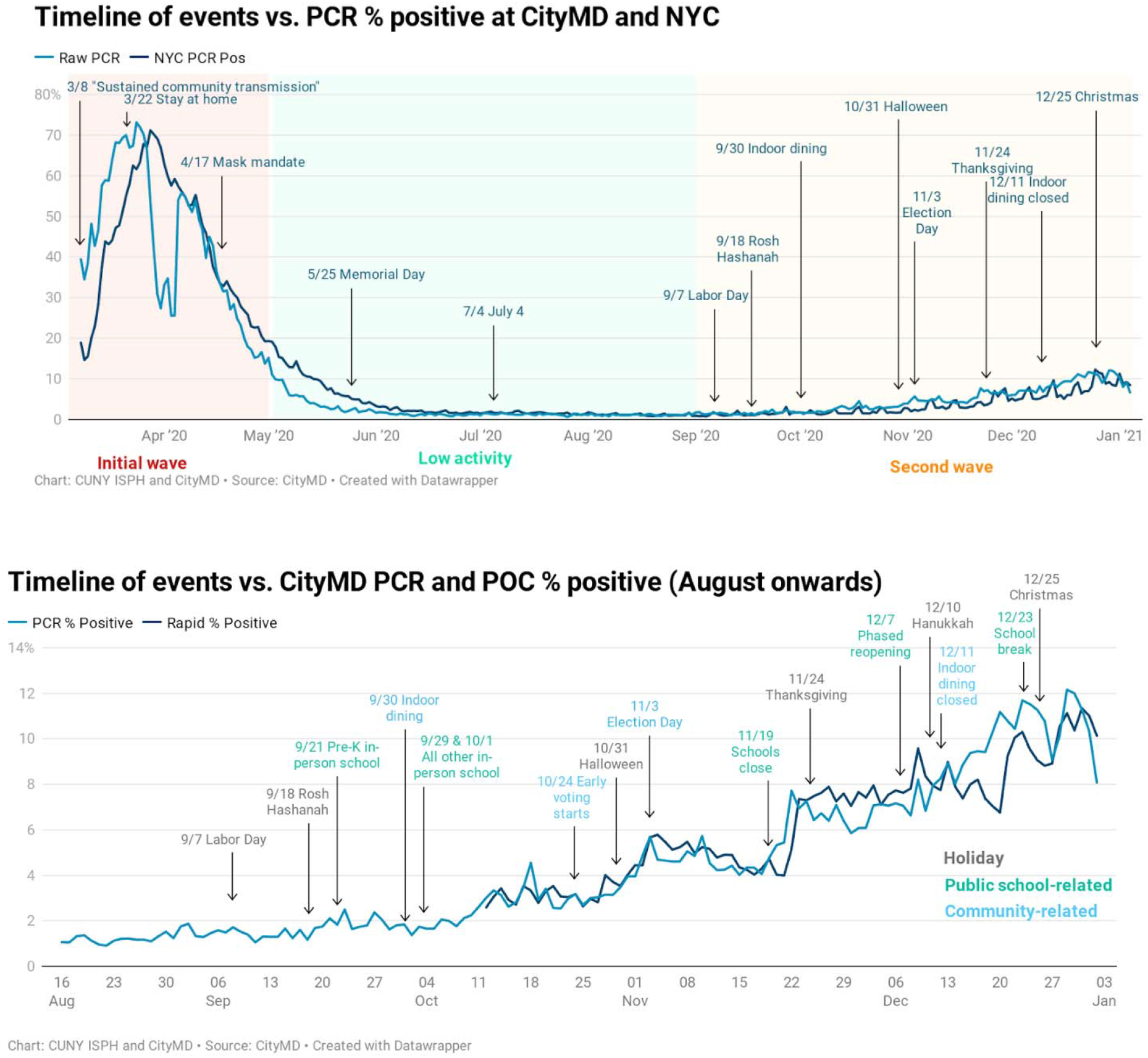
Main events potentially impacting the COVID-19 trends in NYC. Top panel shows PCR percent positivity for PCR tests done at CityMD (light blue) and in NYC (dark blue). The three periods of initial wave, low activity and second wave are shaded in red, blue, and yellow respectively. Bottom panel shows PCR (light blue) and rapid antigen tests (dark blue) positivity rates for CityMD testers focusing on the second wave of the pandemic in NYC. Raw data was used to calculate positivity rates in both plots to be able to visualize the spikes in cases occurring after events that might have facilitated increased in-person contacts. PCR: Polymerase chain reaction. POC: Point-of-care rapid antigen tests.

### Demographic differences in diagnostic testing and positivity

Testing increased for children, adolescents and 20-29 year olds between the two waves (Table 1). About 24% of the patients who received a test at CityMD were Hispanic, 35% were non-Hispanic (NH) White, 9% were NH Black, 7% were Asian, and 0.7% were Native American/Alaskan Indian/Pacific Islanders. Testing trends did not vary by race/ethnicity between the two waves. A higher proportion of those testing were females (55%). Most testers were seen in NYC (67.1%), followed by Long Island (27.1%) and Westchester (5.8%).

Age-specific diagnostic test positivity was highest in the 40-69 year age groups followed by those older than 90 years (Table 2). Those in younger age groups had lower test positivity in the first wave. However, in the second wave, PCR as well as rapid test positivity in 15-19 year olds was high and comparable to those 40-49 and 50-59 years old. PCR positivity was higher in Hispanic, NH Black, and Native American testers compared to NH White and Asian testers (Table 2; Fig 3) in the first wave. Positivity decreased for NH Black (11.3% in wave 1 vs. 5.2% in wave 2) (Table 2) but was still higher compared to NH White testers in wave 2 (4.8%). PCR (3.6%) and rapid test (5.1%) positivity was lowest in Manhattan compared to other regions.

**Figure 3:**
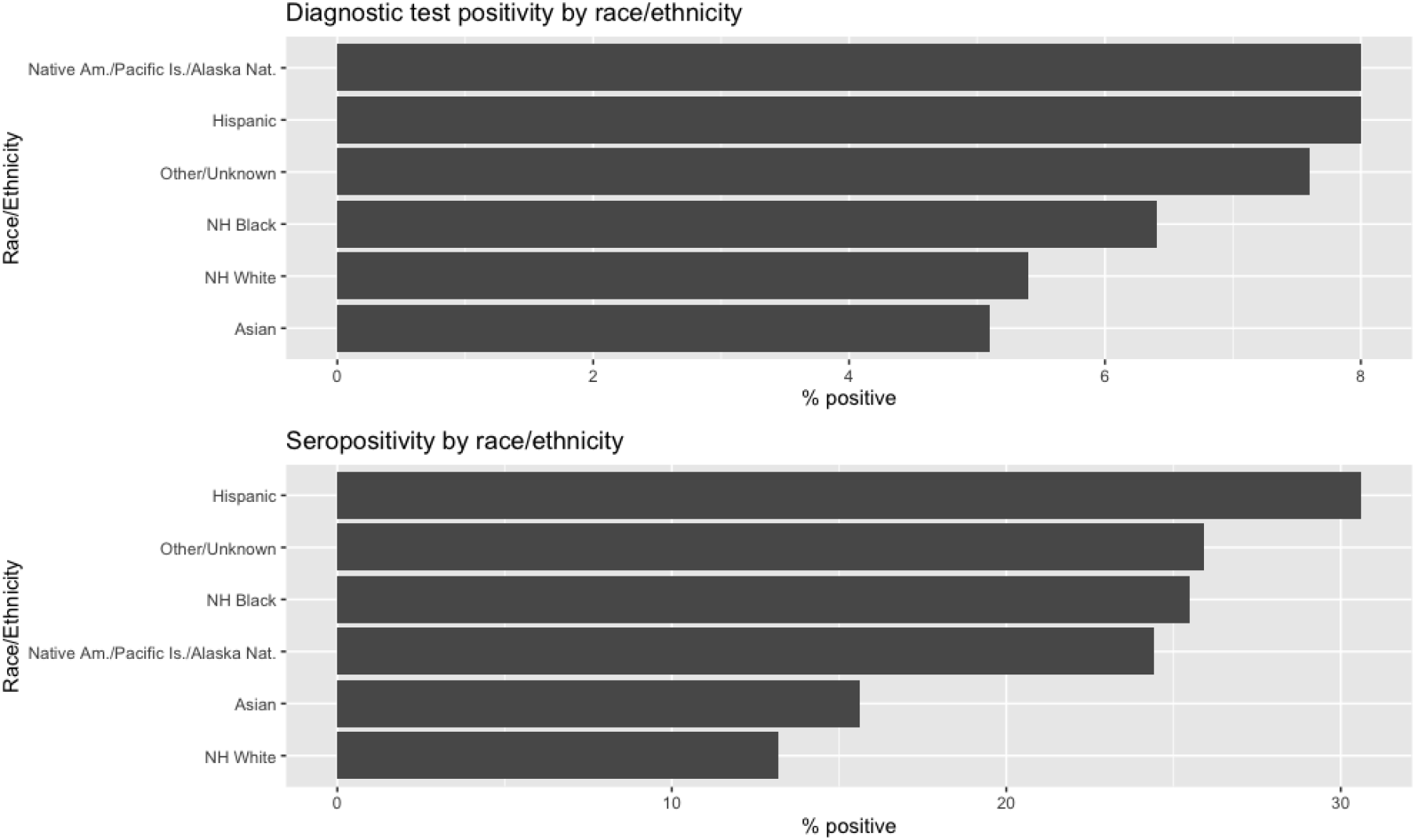
Differences in COVID-19 PCR/rapid antigen test positivity and seropositivity by race/ethnicity. The top panel shows combined PCR and rapid antigen test positivity by race/ethnicity for individuals tested between March 2020 and January 2021. The bottom panel shows antibody positivity by race/ethnicity. Test positivity rates were higher among Native American, Hispanic, and NH Black testers, compared to NH White and Asian testers. NH: Non-Hispanic; Native Am./ Pacific Is./Alaskan Nat.: Native American/ Pacific Islander/Alaskan Native; PCR: Polymerase chain reaction

### Patterns in seropositivity

Overall, and across the entire study period, seropositivity was estimated to be 21.6% (95% CI: 21.5%, 21.7%) among the 693,205 persons receiving at least one antibody test (Table 3). Of the 25,833 individuals who received a diagnostic test prior to a serologic test, 6,788 (26.3%) tested positive by PCR/rapid test; among those 6,788, 5,983 (88.1%) had a positive antibody test. Seropositivity was higher in 5-9 year olds (26.9%) and 10-14 year olds (27.3%) compared to older age groups (Table 3, Fig 4). Seropositivity was also high among individuals over the age of 90 (21.2%), but the number of testers in this age group was relatively small (n=626). Seropositivity estimates were also higher among NH Black (25.4%), Hispanic (30.6%), and Native American testers (24.4%) (Table 3, Fig 3). Residents of the Bronx (31.2%) and Queens (30.5%) had higher seropositivity compared to residents of Brooklyn (19.8%), Staten Island (19.62%), Manhattan (17.0%), Long Island (18.4%) and Metro North (17.8%).

**Figure 4:**
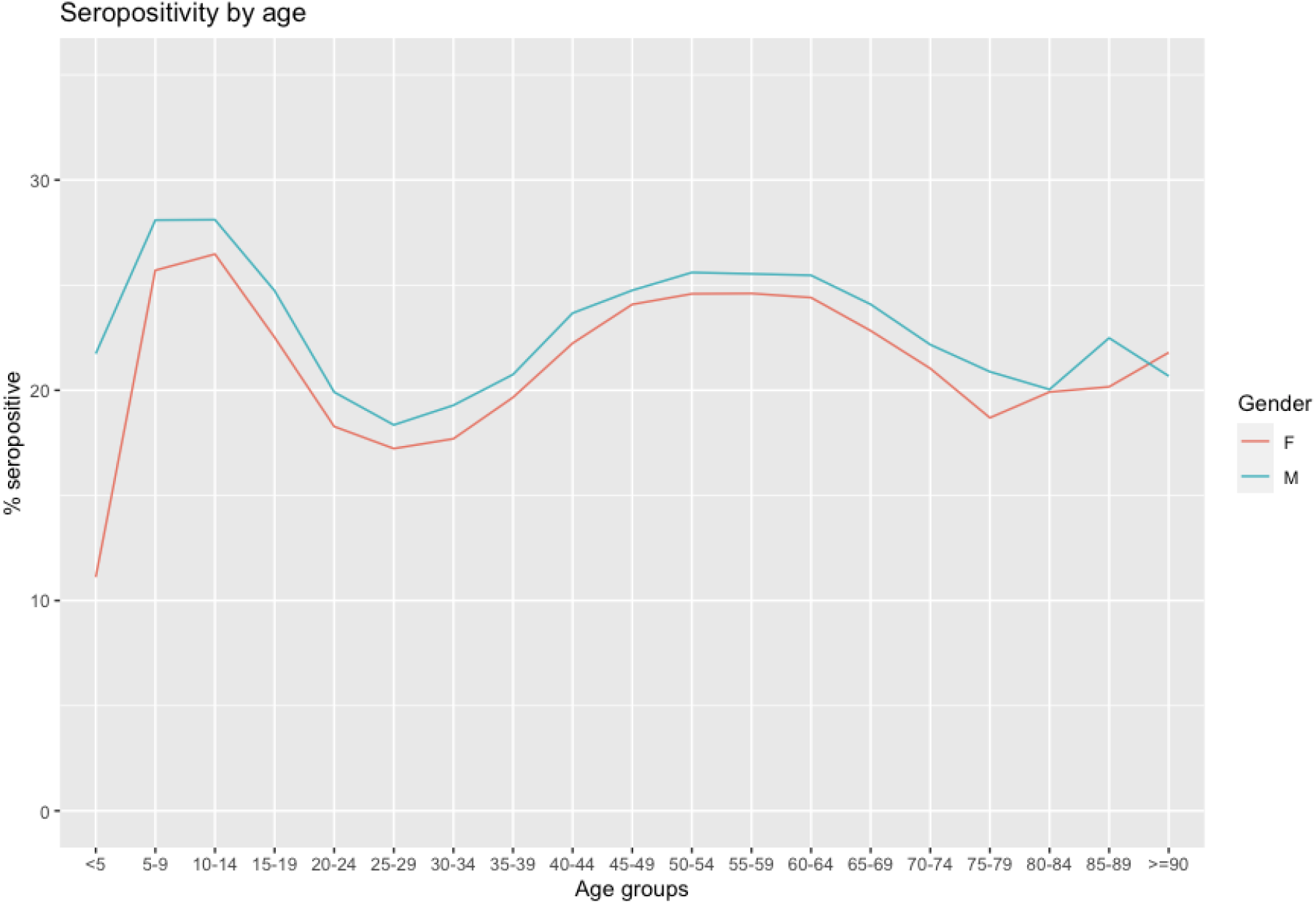
SARS-CoV-2 seropositivity by age and gender between March 2020 and January 2021. Seropositivity for each age group is measured as a proportion of serologic testers who had their first positive serologic test between March 2020 and January 2021. For repeat serologic testers, subsequent positive tests were removed. Red curve is for females and the blue curve is for males. Testers with unknown gender are removed from this plot. F: Female; M: Male.

### False negative and false positive rates for rapid antigen tests

Of the 220,347 rapid antigen tests that were *negative* for SARS-CoV-2 and also tested by confirmatory PCR, 11,824 were PCR positive (false negative rate=5.4%; 95%CI 5.3-5.5). Of these rapid negative-PCR positive patients, 6,646 (56.2%) had COVID symptoms documented in the EMR at the time of testing.

Of the 6,929 rapid antigen tests that were *positive* for SARS-CoV-2 with a confirmatory PCR test, 990 were negative by PCR, giving a false positive rate of 14.3% (95%CI: 13.5-15.1). Of those patients who were rapid positive-PCR negative, 151 (15.2%) were documented as being symptomatic.

### Signs, symptoms, and history of comorbidities

We were able to retrieve vital signs information from the EMR for 75% of patients who were tested. Patients with a positive diagnosis were more likely to present with a fever ≥100.1F compared to those with a negative diagnosis (9% vs. 0.7%). In both waves, a higher proportion of COVID-19 positive individuals had O_2_ levels <95% compared to COVID-19 negative individuals (1.41% in positives vs. 0.3% in negatives in wave 1 and 0.5% in positives and 0.1% in negatives in wave 2). Of these who tested positive by any test, 40,729 (15%) had a documented history of heart disease, 19,303 (7.2%) high cholesterol, 15,679 (5.8%) asthma or COPD, and 10,548 (3.9%) depression and/or anxiety (sTable1).

## Discussion

Using data from a large ambulatory urgent care provider in the New York metropolitan area, we analyzed COVID-19 diagnostic and serologic testing and positivity trends over the course of the pandemic. For NYC only, as of January 8, 2021, CityMD had conducted 27.6% of all PCR COVID-19 tests. Combining PCR and rapid antigen data, CityMD diagnosed roughly 17% of the total SARS-CoV-2 cases in NYC. Seropositivity among CityMD testers in NYC was found to be 22.7%, lower than that of all testers citywide [27%][5], but higher than CityMD patients in surrounding areas of Long Island (18.4%) and Westchester (17.8%). Testing and positivity patterns differed by age, sex, race, and geography, and about 25% of testers were repeat testers. Rapid antigen tests had high sensitivity in this setting. Given the importance of evaluating and testing both symptomatic and asymptomatic cases, epidemic tracking, and controlling pandemic spread, these data highlight the essential role that urgent care providers have played, and continue to play, in serving large numbers of patients during this pandemic. Urgent care providers provide broad access to critical clinical evaluation and testing services and facilitate public health action for ambulatory patients. Additionally, such increased access, early diagnosis and treatment may limit the flow of patients to crowded emergency rooms when alternate settings are more appropriate.

We observed higher infection rates and seropositivity among non-Hispanic Black and Hispanic testers, similar to trends in NYC overall [1] and elsewhere [6,7]. Long-standing barriers and structural inequities in healthcare access might explain these trends [8,9]. Hispanic, non-Hispanic Black and Asian New Yorkers also form a large proportion of essential workers and healthcare workers, further increasing their risk of infection with SARS-CoV-2 [10,11]. It is possible that due to the nature and hours of essential work, there are fewer opportunities and free time to test, making it more likely test only when symptomatic or after a known exposure, resulting in a higher individual and test-level prevalence. Because people of color are more likely to reside in New York neighborhoods that also have higher concentrations of essential workers, and higher levels of household crowding,[12] the virus could have spread more rapidly in these communities early in the pandemic [11]. Therefore, it is critical that evaluation and testing is accessible and without cost barriers (e.g., insurance coverage and patient financial responsibility), especially for these communities.

Seropositivity was higher in 5-19 year old testers compared to 20-44 year olds. Other studies in the US have estimated seropositivity to be lower in younger age groups but they had small samples of children under 18 [13,14]. Low diagnostic testing rates but high seropositivity among children and adolescents suggest probable exposure during the first wave (e.g., while in school during high levels of community spread prior to lockdown) but were not tested because of testing availability, or due to having milder symptoms or being asymptomatic [15]. This could have implications for transmission from younger children to older, more vulnerable age groups [16].

The overall prevalence of antibodies in this cohort of SARS-CoV-2 testers was 21.6%, suggesting that a majority of individuals remain susceptible. Moreover, recent studies suggest waning of SARS-CoV-2 antibodies to both nucleocapsid and spike proteins [17] which could mean that the true proportion of testers who were exposed to SARS-CoV-2 is higher. Population representative surveys can provide better estimates of the true seropositivity and cumulative incidence of SARS-CoV-2, and thus better estimate the number and sociodemographic characteristics of residents who remain susceptible at a given time.

With the introduction of SARS-CoV-2 rapid tests with faster turnaround times, testing has become even more convenient and has greatly improved the ability to confirm active symptomatic infection and screen for asymptomatic and pre-symptomatic infections. Rapid tests were highly sensitive (∼94%) in this urgent care setting. Rapid tests are a useful diagnostic tool for quickly identifying infectious persons because PCR tests, though more sensitive, can have long turnaround times and be positive well after the end of the infectious period [3]. Rapid tests are also less invasive than PCR tests, which makes them more popular among testers, and have the potential to increase testing uptake broadly [18,19]. Widespread availability of rapid tests, promptly followed by self-isolation has great potential to mitigate community spread.

Using longitudinal EMR data, we described repeat testing patterns, an aspect of testing during the pandemic that has not been well-characterized yet. About 25% of the patients were repeat testers and close to 60,000 received four or more diagnostic tests. These individuals are likely following NYC Department of Health and Mental Hygiene’s (NYC DOHMH) recommendations of frequent testing for those who are healthcare, essential workers, and who cannot work from home [20] to ensure cases are diagnosed early and that public health action follows (e.g., isolation). Easy access to testing is critical to adhering to employer policies; availability of rapid tests at CityMD likely further enabled patients to test regularly [18].

We found that 3,604 individuals presented at CityMD clinics for subsequent testing within 10 days of a prior positive test (i.e., during their isolation period) and received a second positive test. A majority of these tests (n=3,225, 89.5%) happened post-October 2020 and were rapid tests. It is possible that these individuals were testing as required by employers to be able to return to work or to prepare for holiday gatherings. However, when individuals test while still infectious during subsequent visits, they could potentially expose clinic staff and other testers. While testing frequently is encouraged in general, CDC recommendations as of October 21, 2020 state that those with a positive test should isolate and not test again for at least 3 months [21], because a PCR test can be positive for ∼90 days post infection. Governments should consider clearer messaging around repeat testing with both PCR and rapid tests after an initial positive test focused on the possibility of transmission at the testing site during the infectious period and the importance of self-isolation for the entire isolation period. Workplaces that require employees to get negative tests before resuming work should evaluate policies to ensure they do not conflict with public health recommendations.

The testing and positivity trends over time in our study population closely mirror overall NYC population-level testing trends reported by the NYC DOHMH [22]. Detailed self-reported information on race and ethnicity allowed us to examine testing frequency and positivity rates by categories that are not usually available. Our results suggest that the OMB categories for race/ethnicity can mask wide variability and exceedingly high SARS-CoV-2 prevalence in some instances. For example, the overall diagnostic positivity and seroprevalence among Hispanic testers was 8% and 29.4%, respectively. However, within Hispanic ethnic groups, these proportions ranged from 4%-16% and 14%-52%, respectively (sFig3, sFig4). Health care providers and public health jurisdictions should endeavor to collect more complete and accurate self-reported data on race/ethnicity, as these data improve our understanding of disease risk and remain of considerable epidemiologic importance for most health conditions [8].

Our study has limitations. Our data only includes individuals who sought a COVID-19 test. Patients who seek care at CityMD are not representative of all persons testing for SARS-CoV-2 in the NY metropolitan area or the general population. Because CityMD is an ambulatory care provider, we did not have information on clinical evaluation and testing outside of CityMD, subsequent development of severe disease, hospitalization, or death after the visit. Typical COVID-19 symptoms were not captured in a standardized form in the EMR and could not be analyzed.

In summary, our results highlight the vital role that urgent care providers play in evaluating, diagnosing and treating substantial numbers and proportions of patients for COVID-19, and in triggering self-isolation, contact tracing and helping to limit onward spread especially in population-dense, urban epicenters. CityMD may have limited the flow of less severe patients to emergency departments of hospitals, which was of critical importance during periods of surge. Also, early identification of cases in urgent care centers results in early interventions which could lead to lower morbidity. Future pandemic preparedness plans should leverage urgent care providers for a multitude of critical implementation roles with the potential to improve individual and public health outcomes.

## Data Availability

The data used in this manuscript is not publicly available.

