## Supplemental Materials for "The emergence, surge and subsequent wave of the SARS-CoV-2 pandemic in New York metropolitan area: The view from a major region-wide urgent care provider"

**Supplementary material**

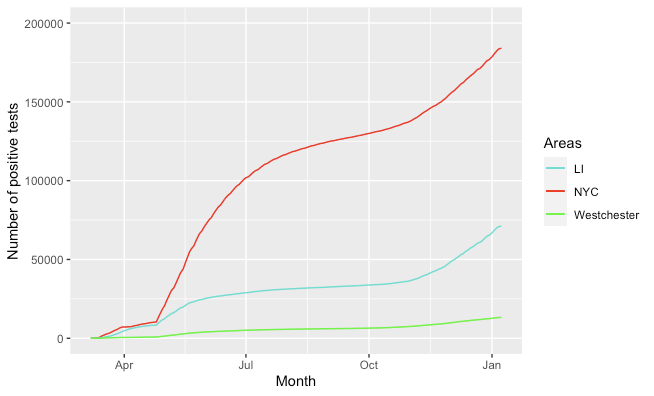

**sFig 1: Cumulative incidence of COVID in among NYC residents testing at CityMD**

Only the first positive result is included while estimating cumulative incidence. Curve in red is for NYC, blue for Long Island, and green for Westchester. Most cases diagnosed were from NYC. Cases are defined as individuals who received their first positive test result when tested with either a PCR, rapid antigen, or serologic test. LI: Long Island; NYC: New York City.

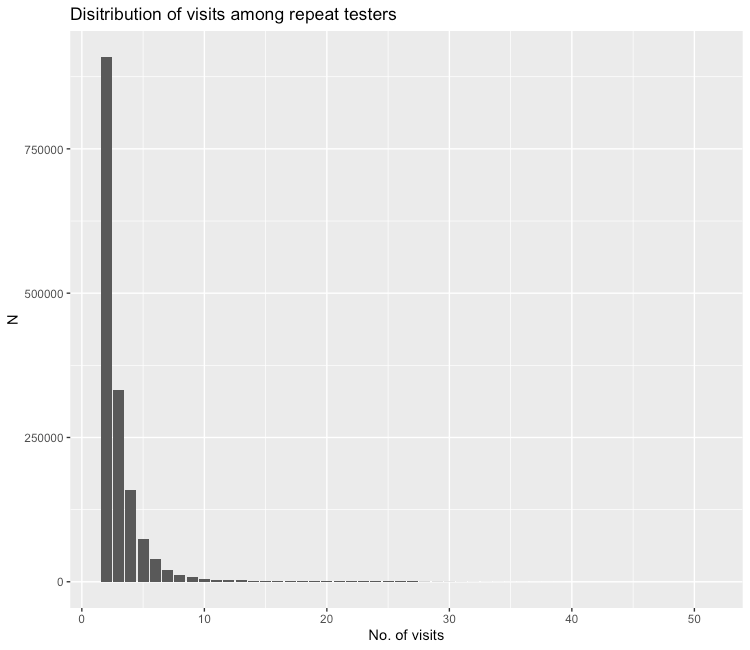

**sFig2: Distribution of number of visits among repeat testers**

A majority of the repeat testers had 2 tests of any type. The maximum number of tests performed by a single tester was 51.

**Diagnostic test positivity and seropositivity by self-reported race and ethnicity**

Self-reported race in the CityMD EMR included specific Native American tribes and country-level nationalities. First, we categorized reported races into NH White, NH Black, Asian, Native American/Pacific Islander/Alaska Native, and Other/Unknown based on the values reported in the “Race” variable using OMB guidelines. For patients missing a value for race, their responses recorded for the “Ethnicity” variable were used to assign ethnicity. Any patient who reported “Hispanic” race  was classified as having “Hispanic” OMB race/ethnicity category, resulting in the following categories: non-Hispanic White, non-Hispanic Black, Asian, Native American/Pacific Islander/Alaska Native, Hispanic, and other/unknown. Patients who had both race and ethnicity missing were classified as having “other/unknown” race/ethnicity.

Patients reported 896 different drop-down categories for race, and 60 of these categories were listed by more than 1,000 individuals. Patients reported 44 different categories for ethnicity, of which 36 were listed by more than 1,000 persons. We present here estimates of SARS-CoV-2 seropositivity and diagnostic test positivity for the different self-reported racial and ethnic groups with more than 1000 people in them.

 Patients that identified as Spanish American Indian, Central American Indian, and Bangladeshi under race had ~15% infection rate, while those who identified as Guatemalan, Honduran, and Salvadoran under ethnicity had ~13% infection rates. Spanish American Indians, Central American Indians, Bangladeshis, and Mexican American Indians also high seropositivity compared to other races (>30%), while patients identifying as Mexican and Ecuadorian ethnicities over reported 40% seropositivity rate. On the other hand, patients who identified as Japanese, Korean, Vietnamese, Chinese, Chilean, Argentinian, Cuban, and Andalusian as their race or ethnicity had low infection and seropositivity rates.

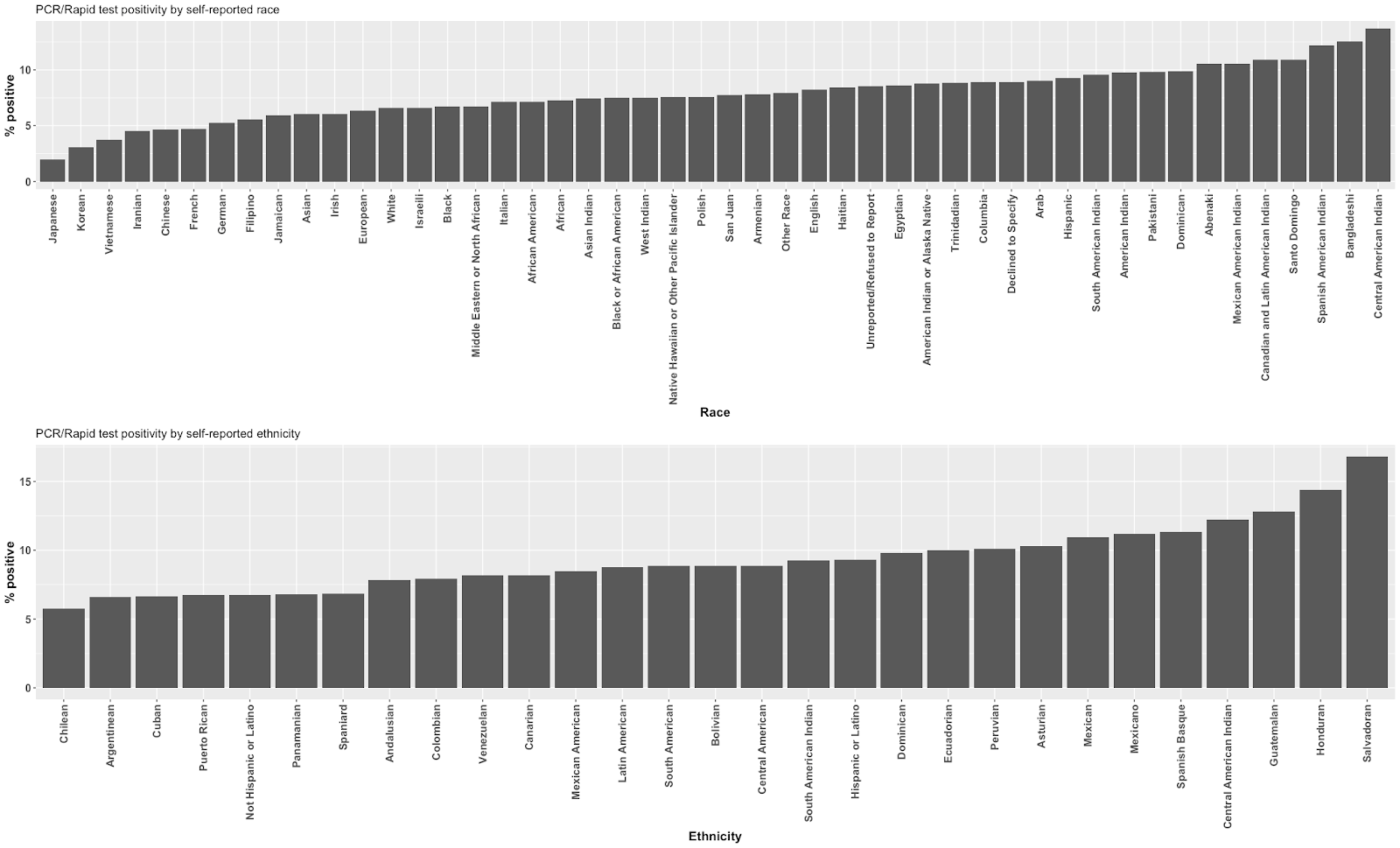

**sFig3: Percent positivity of diagnostic tests by self-reported race and ethnicity**

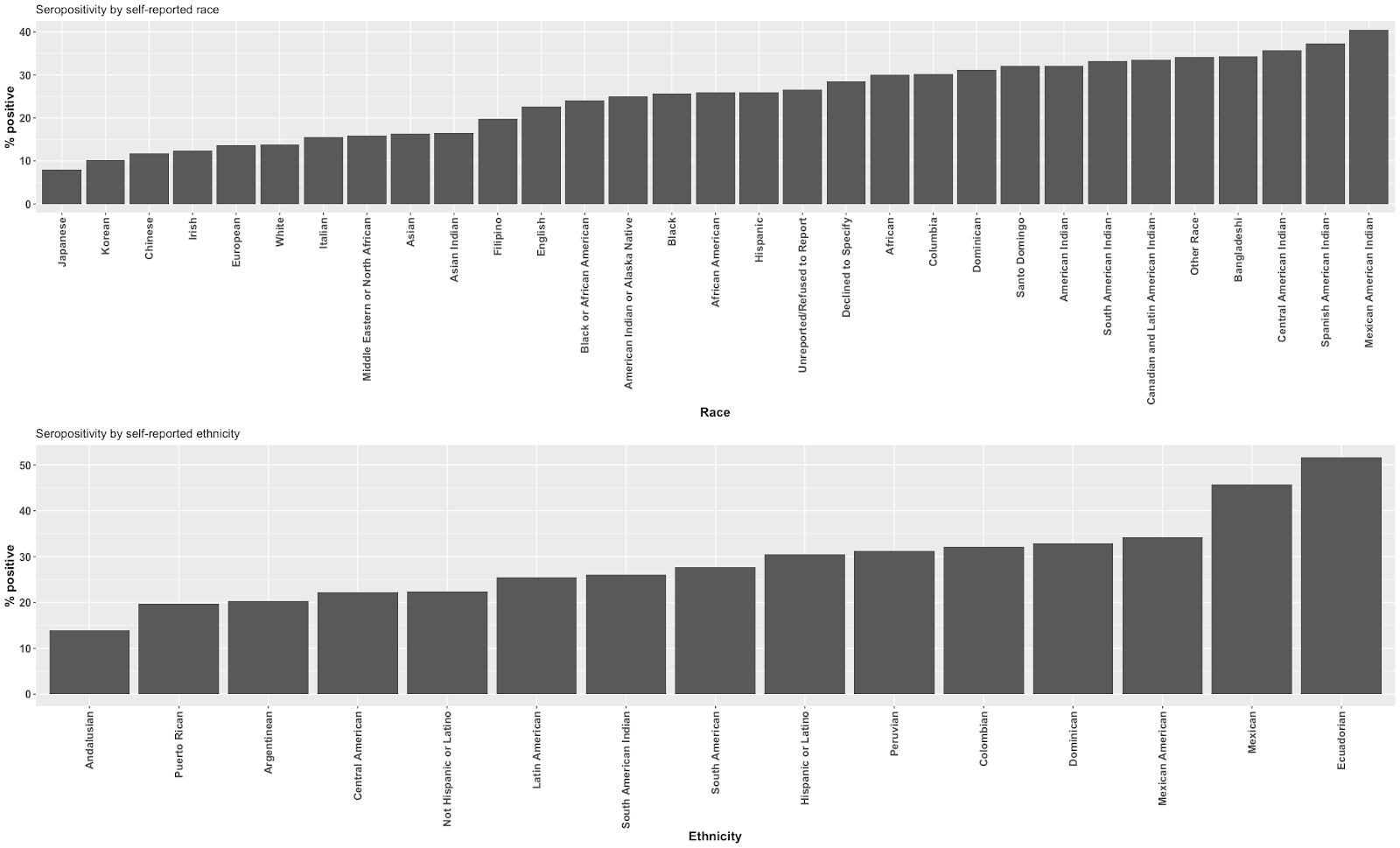
**sFig4: Percent positivity of diagnostic tests by self-reported race and ethnicity**
